# From Spreadsheets and Bespoke Models to Enterprise Data Warehouses: GPT-enabled Clinical Data Ingestion into i2b2

**DOI:** 10.1101/2025.04.17.25325962

**Authors:** Taowei David Wang, Shawn N. Murphy, Victor M. Castro, Jeffrey G. Klann

**Author notes:** **(corresponding author) Corresponding author information:** 45 Irving St., Medford, MA 02155; ph.

## Abstract

**Objective:** Clinical and phenotypic data available to researchers are often found in spreadsheets or bespoke data models. Bridging these to enterprise data warehouses would enable sophisticated analytics and cohort discovery for users of platforms like NHGRI’s Genomic Data Science Analysis, Visualization, and Informatics Lab-space (AnVlL). We combine data mapping methodologies, biomedical ontologies, and large language models (LLMs) to load these data into Informatics for Integrating Biology and the Bedside (i2b2), making them available to AnVIL users.

**Materials and Methods:** We developed few-shot prompts for ChatGPT-4o to generate Python scripts that facilitate the extract, transform, and load (ETL) process into i2b2. The scripts first convert a designated data dictionary (in various formats) into an intermediate common format, and then into an i2b2 ontology. Finally, the original data file is converted into i2b2 facts, using standard ontologies hosted by the National Center for Biomedical Ontology (NCBO).

**Results:** ChatGPT-4o correctly produced Python code to facilitate ETL. We converted phenotype data from three synthetic datasets from three disparate data models available in AnVIL. Our prompts generated scripts which successfully converted data on 3,458 fake patients, making it queryable in i2b2.

**Discussion:** For a few datasets, iterative prompt refinement might reduce ETL efficiency gains. However, prompt reuse significantly reduces incremental effort for additional data models. At scale, we anticipate our pipeline offers substantial time savings, which could transform future ETL workflows.

**Conclusion:** We developed an LLM-powered ETL pipeline to convert disparate datasets into i2b2 format, enabling advanced analytics and cohort discovery across heterogeneous data models.

## Background and Significance

In less than three years since OpenAI’s launched ChatGPT-3, making Large Language Models (LLMs) accessible to the general public, LLM applications to medicine are emerging on an almost daily basis.[1] LLMs are able to assist with chart review, diagnosis, and query design. Recently, experts have suggested that, more broadly than medicine, LLMs will disrupt data management, changing our approach to “a number of hard database problems.” [2] Here, we present a novel approach to the Extract, Transform, and Load (ETL) problem that couples LLMs with traditional knowledge base approaches.

Clinical data collected for specific research studies is often stored in spreadsheets, with many columns, one for each variable collected. The variables tend to be granular and study-specific, such as “Date of first diagnosis” or “Affected Status”. Frequently the data are not coded into standard terminologies or unusual terminologies are used.

To bring spreadsheet data to clinical data warehousing platforms, the data must be ETL’d into platform-compatible structures. Typically, this is a time-consuming per-project programming effort for every custom spreadsheet. If this process could be simplified and streamlined, it would allow researchers with spreadsheet-style data to utilize the graphical cohort discovery and analysis tools in platforms like Informatics for Integrating Biology and the Bedside (i2b2)[3] and Observational Health Data Sciences and Informatics (OHDSI).[4] It would also enable loading hybrid-model datasets like Electronic Medical Records and Genomics (eMERGE)[5] into Common Data Models (CDMs).

There is significant clinical data stored in these spreadsheets and bespoke data models, as can be seen through a variety of lenses. First, patient registries collect data to study specific conditions, often combining EHR and patient survey data. Registries are prevalent and growing. As of 2020, over 600 registries with data on over a half-million patients total were registered on ClinicalTrials.gov.[6] Each of them uses registry systems with bespoke data models, limiting cross-system analysis. dbGaP (Database of Genotypes and Phenotypes) datasets of genetic and health-related data are available to researchers under controlled access. dbGap reports 5,764 datasets with phenotype tables, and 794 include the keyword “phenotype,” including prominent datasets from well-known programs, such as the Trans-Omics for Precision Medicine (TOPMed) program, the Genotype-Tissue Expression (GTEx) project, and the Undiagnosed Diseases Network (UDN).[7] Each of these defines a custom data dictionary (some are similar, but none identical). Secondly, researchers have found significant presence of Excel files submitted as supplemental data for genomics publications in PubMed between 2016 to 2020 (32,000 files for 166,000 papers).[8] This is an underestimate, because spreadsheet-compatible plain text formats (e.g. .CSV, .TSV) are not accounted for. Finally, given the vast number of healthcare facilities and clinical studies globally, it is easy to extrapolate that the number of spreadsheet-like datasets are in the hundreds of thousands, reflecting the decentralized and heterogeneous nature of data collection in the industry.

Significant data integration efforts have been made, often requiring painstaking work and investments of millions of dollars. BioDataCatalyst is a cloud computing platform designed to facilitate analysis on biomedical datasets. It supports the Patient Information Commons and Standard Unification of Data Elements (PIC-SURE), which provides integrated analysis tools on several high-impact data sources, including TopMED and BioLINCC.[9] It is infeasible to replicate this investment on thousands of different spreadsheet formats.

This study’s focus is on the NHGRI Genomic Data Science Analysis, Visualization, and Informatics Lab-space (AnVlL). AnVIL is a cloud-based platform supporting 15,000 clinical genomic researchers through a scalable, shared environment with powerful analysis tools and over 5 petabytes of data from various consortia (including dbGAP).[10] The recently-funded AnVIL Clinical Environment for Innovation and Translation (ACE-IT) aims to enable AnVIL to support clinical researchers with clinical data tools and clinically-focused genomics tools, such as polygenic risk score calculators and machine-learning diagnostic tools. The clinical data in AnVIL datasets fall into this semi-structured spreadsheet-like format. To utilize it in ACE-IT, the data must be standardized into a CDM and a structured coding system.

### i2b2

i2b2 is a well-established, open-source clinical data warehousing and analytics platform in use at over 300 locations worldwide.[3] Over 15 years, i2b2 has evolved from a single-site cohort identification tool into an enterprise-ready, network-enabled analytics platform with a diverse open-science community. Its emphasis on flexibility makes it an ideal data integration platform. It can represent non-standard healthcare data (e.g., social determinants of health, survey instruments, and patient-reported outcomes). In addition to the i2b2 data model, the software also supports Observational Medical Outcomes Partnership (OMOP) Common Data Model. i2b2’s primary interface is a graphical cohort-finding tool, which can be used as a self-service portal by researchers to perform analytics and refine a cohort.

In i2b2, patient data are stored as “facts” in a long fact table. For example, one encounter might generate many facts: e.g., diagnosis codes, laboratory tests, and vital signs. Additional information on e.g., patients and visits are stored in dimension tables. The meaning of codes and values are defined in a separate table, called an “i2b2 ontology,” which defines a hierarchy of nodes, with metadata about each. For example, the ICD-10-CM ontology begins with high level groupers like “Neurological Disease” followed by several levels of subcategories, like “Nerve, nerve root, and plexus disorders,” until finally leaf nodes like “Bell’s Palsy” are found. Queries can be made on folders or leaves.

### LLM code writing

Large Language Models (LLMs) are becoming valuable assistants for coding tasks. Initially developed for natural language tasks such as translation of human languages, LLMs have shown surprising effectiveness in translating human-readable text into computer languages as well. In fact, LLM-assisted programming is an area of active research, as is benchmarking LLM code-generation correctness. [11–13] Microsoft’s GitHub Copilot, for instance, is a tool designed to support programmers by leveraging OpenAI’s Codex, which was trained on a vast collection of publicly available code, including GitHub repositories. [14] Copilot was shown to improve time efficiency up to 36% in real world software engineering projects. [15] While Copilot is primarily aimed at experienced coders, LLMs have the potential to make programming more accessible to non-programmers because of their ability to take descriptions of coding tasks in natural language and output functional code. Prompt engineering, the process of crafting LLM input to produce desired outputs, plays a crucial role in this context. [16] While a simple programming task can be described to an LLM in a few sentences, a complex algorithm or data transformation is much more involved. Due to the length of such prompts and LLM limitations, this requires careful refinement of prompts, with sufficient reminders throughout of constraints described early-on. Emerging applications are beginning to demonstrate the potential of this approach.[17]

## Objective

We seek to automate conversion of spreadsheet data into i2b2 format by using carefully engineered LLM prompts combined with sample data and data dictionaries to generate Python programs. With ChatGPT-4o, we apply this to the conversion of three well-defined phenotypic datasets available to AnVIL users, as a blueprint and prompt library for additional i2b2 conversion. These datasets are synthetic versions of data from the Centers for Mendelian Genomics (CMG), Genomics Research to Elucidate the Genetics of Rare Disease (GREGoR), and eMerge III (Figure 1).

**Figure 1.**
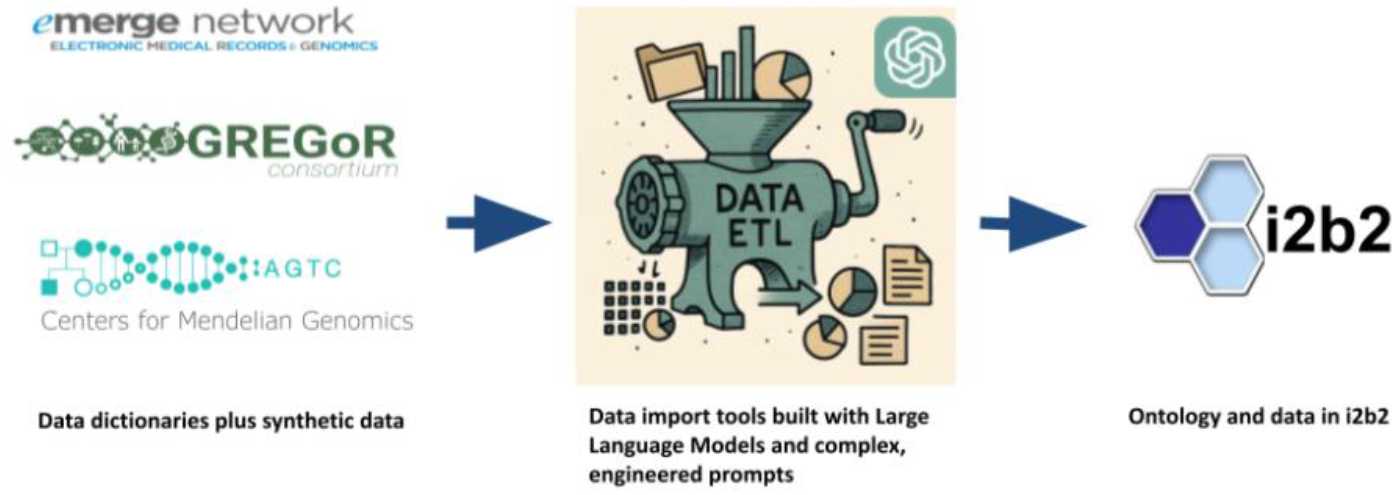
Conceptual methodology to extract-transform-load heterogeneous data dictionaries and accompanying data into i2b2 through a LLM-generated pipeline. (The “data-grinder” - center - was generated by ChatGPT.)

## Methods

We chose three data sources for conversion, based on their familiarity to AnVIL users. Data dictionaries were obtained from public websites, and collaborators at Vanderbilt University Medical Center generated sample synthetic data tables with characteristics like a real study, as described below.

- The GREGoR Consortium (Genomics Research to Elucidate the Genetics of Rare diseases) develops and applies approaches to discover the cause of currently unexplained rare genetic disorders. Phenotype and participant data dictionaries were acquired from the Consortium’s public repository,[18] and a 94-patient synthetic dataset was generated.
- The Centers for Mendelian Genomics use next-generation sequencing and computational approaches to discover the genes and variants that underlie Mendelian disorders. We acquired the CMG Subject data dictionary as a CSV spreadsheet and sample data on 1000 synthetic patients. The CMG Subject table was derived from the data model schema,[19] and the original data come from the AnVIL CMG-Yale workspace. [20]
- The eMERGE Network is a large-scale research project that aims to investigate genetic and environmental factors that contribute to various health conditions. We acquired the Subject/Phenotype data dictionary as an XML description along with sample data on 2364 synthetic patients from the eMERGE III Human Reference Consortium study. This includes case/control status for a wide array of phenotypes.

Our source data consisted of data dictionaries and many-column CSV synthetic data files. We designed a pipeline for conversion to i2b2 using three major components (Figure 2). The first step is the translation of the data dictionaries into a common format. The next two steps are generating an i2b2 ontology and a fact table. The ontology defines all possible fact types and their names and codes, arranged in a hierarchical structure. The fact table is a pivot of the many spreadsheet columns into the one-row-per-fact i2b2 format, with the values translated into the codes and data defined by the ontology. A more detailed description is below, followed by our LLM-powered approach to implement the methodologies.

**Figure 2.**
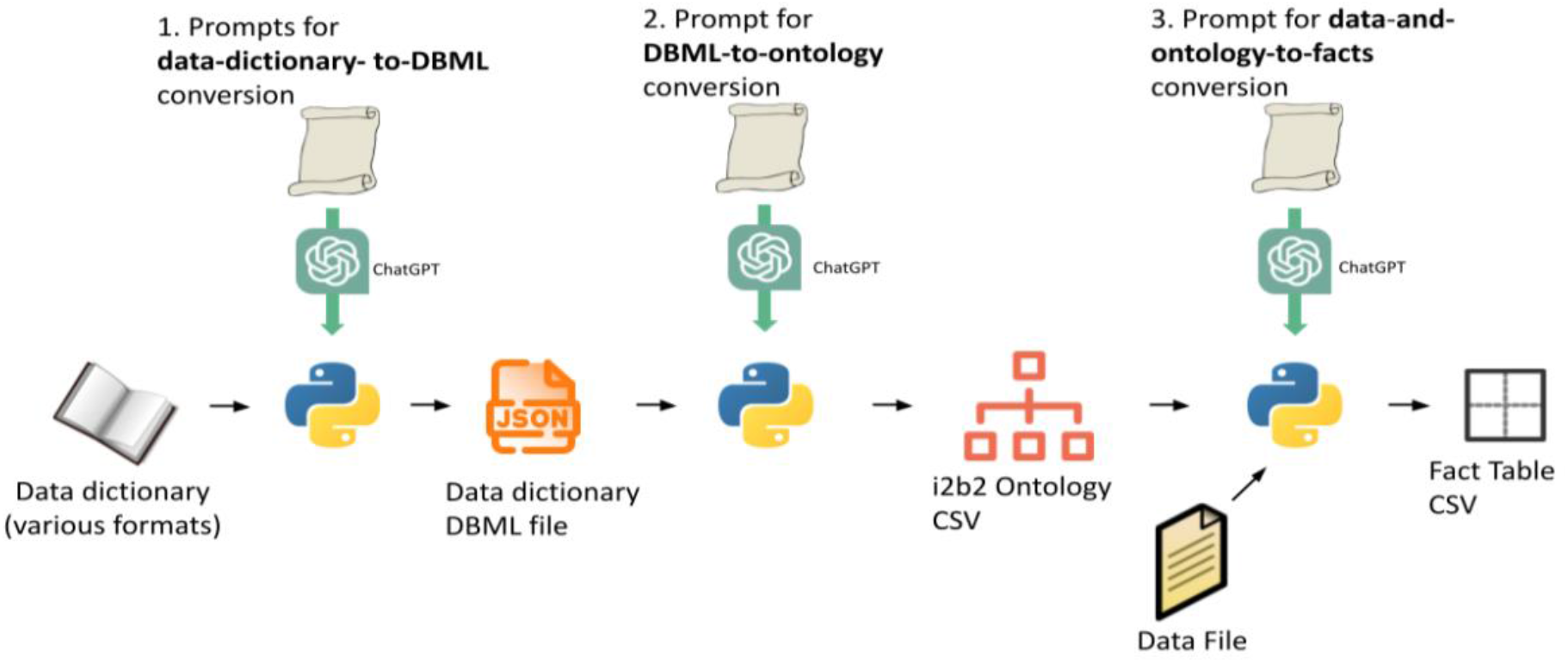
ChatGPT generates Python programs from our detailed prompts (which include e.g., technical documentation and sample data). Python programs convert data and data dictionaries into i2b2 facts and ontologies.

### Data conversion methodology

We harmonized to a common data dictionary format based on GREGoR’s approach. GREGoR’s data dictionary uses a custom JSON format derived from the Database Markup Language (DBML). This structured format allows expression of all three data dictionaries. The DBML structure defines *tables* with *columns*, corresponding to CSV data files. Each column is defined as text, numeric, or an enumerated list. This can be visualized as a shallow ontology hierarchy, with leaves for text/numeric and folders for enumerations (with enumeration as leaves underneath them). A special case occurs when a column refers to a standard terminology, like ‘HPO’. In all three datasets, columns in this format have text values in the format “<terminology>:<value>“, e.g., “HP:0030786” is the Human Phenotype Ontology code for *photopsia*. All three data dictionaries could be converted to this format, because they all used a simple category/enumerated-value hierarchy.

To generate an i2b2 ontology from a structured DBML JSON specification, we developed the following approach. Each JSON field object is translated into either an ontology leaf node or a folder and several leaf nodes, depending on data type. Enumerated fields are expanded into a parent folder with child leaf nodes for each possible enumerated value. The ontology hierarchy, defined by a backslash-separated string of ontology node names, is constructed by combining table and field names with enumeration values, using underscores for spaces and ensuring all paths end with a trailing backslash. Node attributes such as C_NAME, C_BASECODE, and C_VISUALATTRIBUTES are assigned based on naming rules and content. Additional metadata, including comments, tooltips, and system codes, is extracted from the JSON and included in the output.

To convert spreadsheet data into an i2b2-compatible fact table, each row of the input CSV file is treated as a single encounter. Patient identifiers are first mapped to i2b2 pseudoidentifiers using an external patient mapping table. A unique encounter number is then assigned to each row, along with a start date derived either from a user-specified column or a default value. For each data column in the row, the corresponding ontology path is located. If a cell value is formatted as `<code_system>:<code>`, it is interpreted directly as a concept code, bypassing additional ontology logic, which allows integration of external terminologies without requiring full ontology expansion (see “Standard bioportal ontologies” below). If the column maps to a leaf node in the ontology, a fact is created using the predefined code along with the cell’s text or numeric value. If the column corresponds to a folder node, the data value is used to locate the matching leaf node within that folder, and a fact is generated using the associated code, without an additional value field.

### LLM-assisted data conversion

To implement these tasks, we employed a methodology utilizing Large Language Models (LLMs) to assist in writing data integration programs. This process involved developing detailed prompts describing the input and desired output, which resulted in Python programs to process the source data into i2b2 tables and ontologies. The prompts serve as templates that can be modified for specific input formats.

We developed the following methodology for prompt engineering.

1. Write an initial prompt, stating that the objective is to output a Python program to perform the data conversion task (e.g., DBML JSON data dictionary to i2b2 ontology CSV). These elements are included:
  a. A detailed description of the steps needed to complete the task.
  b. Portions of technical documentation that are necessary to develop a solution (e.g., i2b2 ontology table definition).
  c. Notes on important things to pay attention to (e.g., make sure C_FULLNAME is never null).
2. Iteratively refine the prompt by running the Python program, examining the output, and interactively making suggestions for program changes to ChatGPT.
3. When the output satisfies expectations, ask ChatGPT to generate a revised prompt that integrates the changes. Edit the resulting prompt and then feed it to a new ChatGPT session, to test whether the one-shot output of the Python program is correct.
4. Save the prompt and code.

### Standard bioportal ontology conversion

Some source data from GREGoR, CMG, and eMERGE are already encoded in standard ontologies. In this case, we instruct the LLM to retain the standard ontology codes during fact conversion. We then separately convert the standard ontologies hosted in the National Center for Biomedical Ontology (NCBO) Bioportal into i2b2.[21] We developed an updated version of the i2b2 NCBO Extractor tool to programmatically download and extract the ontologies through NCBO’s REST API.[22,23] It includes sample Python scripts, generated with assistance from ChatGPT, to aid users in database integration.

The i2b2 NCBO Extractor comprises two main commands: *ExtractAll* and *ProcessAll*. The *ExtractAll* command downloads an ontology, specified by its unique ID, from BioPortal and extracts the concepts into a pipe-delimited staging file. Each concept contains reference to its parent concepts and requisite i2b2 properties, facilitating import into a staging table in the database (Figure 3’s first two steps).

**Figure 3.**
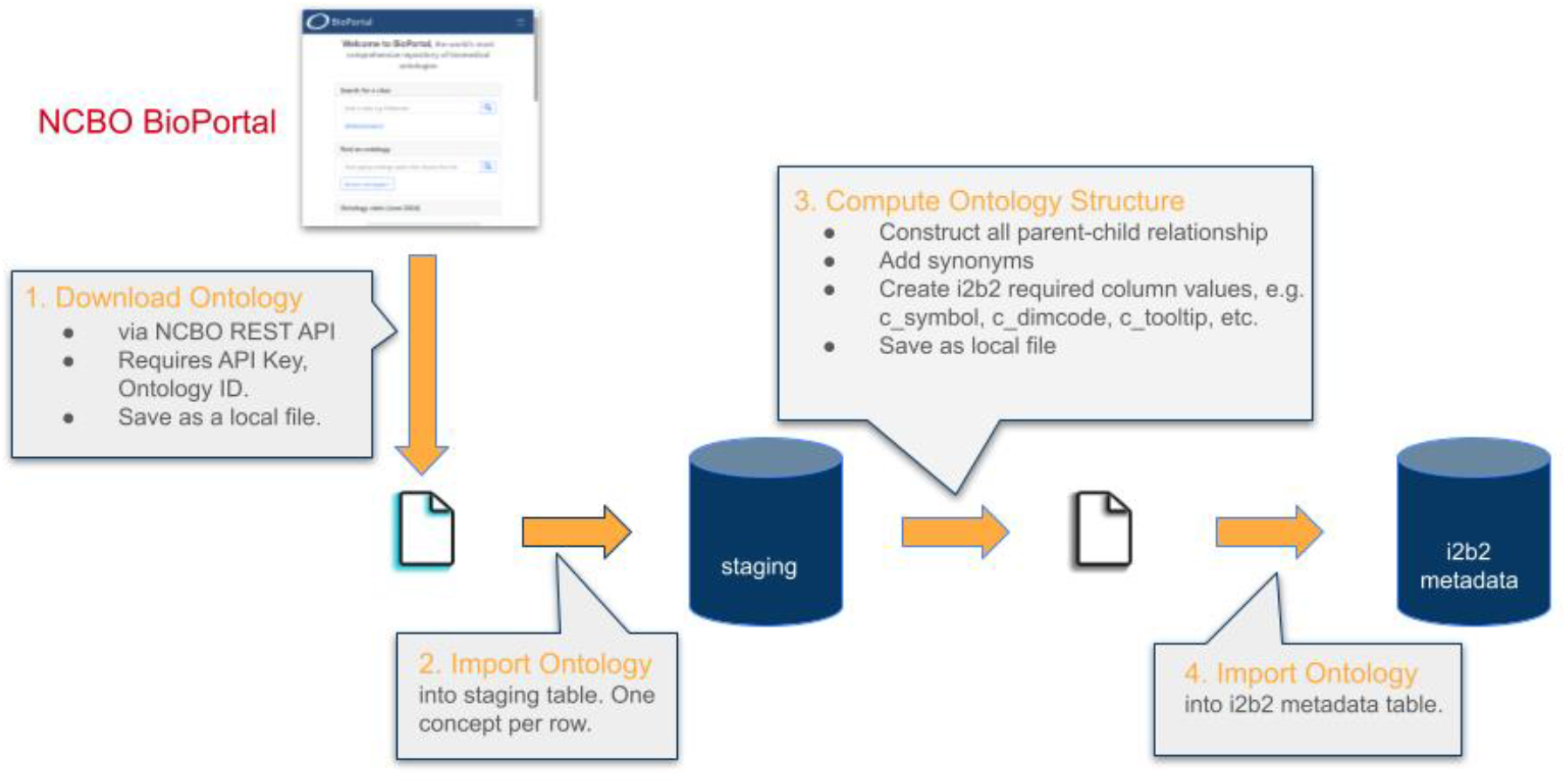
Illustration of the NCBO Extractor workflow, from downloading an ontology from NCBO BioPortal to loading the final, converted ontology into an i2b2 instance.

We subsequently run the *ProcessAll* command to produce a final metadata file for loading into the production i2b2 database. The command requires database connection information and utilizes SQL queries to efficiently construct the ontology hierarchy from the staging table, ensuring all concepts are accounted for. The resulting output is a pipe-delimited metadata file containing all concepts and i2b2-required columns, ready for import into an i2b2 instance (Figure 3’s last two steps).

## Results

We developed prompts that generate Python code to convert data dictionaries to i2b2 ontologies and data files to the i2b2 fact table (Figure 2), along with an ancillary prompt for mapping patient pseudo-ids. All prompts are available at https://github.com/jklann/LLMETL.

### Prompt: DBML data dictionary format

To translate local data dictionaries into GREGoR DBML JSON format, we developed one-off prompts to generate Python code. Given their straightforward nature, we challenged ChatGPT’s capabilities by providing under- specified prompts that were mostly example inputs and outputs.

For CMG (CSV to DBML), we provided the data dictionary’s column headers, two sample rows, and a detailed description of the DBML format. After successfully translating these sample rows, we asked ChatGPT to generate a Python program to do the full translation. Through iterative refinement (necessary due to our under-specification), the final Python program operated correctly. We asked ChatGPT to generate an updated prompt based on the refinements, from which ChatGPT regenerated the Python program without error.

For eMerge III (XML to DBML), we provided ChatGPT the eMerge III XML file and a portion of the GREGoR DBML file (including an enumerated value example) but minimal additional instruction. The model produced a plausible DBML translation, so we asked it to generate a prompt for creating a Python converter. The resulting script produced correct output without modification, likely due to the input being limited to enumerated-value fields.

### Prompt: i2b2 ontologies

To convert DBML data dictionaries into the i2b2 ontology data structure, we developed a prompt explaining the approach described in the Methods. Following this, we included a technical description of the desired program and its inputs and outputs. Key elements included: a set of rules for mapping a DBML column to a leaf node and doing Metadata XML generation; rules for creating a hierarchical structure for enumerations; and, a convention for creating i2b2 codes.The prompt also included significant background information: A sample DBML file, detailed descriptions of the i2b2 ontology format (including excerpts from the i2b2 documentation), and details of the Metadata XML used to define i2b2 pop-ups that allow text or numeric input. This prompt required several rounds of iterative development, due to the complexity of the task and challenges of LLMs (see Discussion).

Table 1 shows an example of the main program’s task, specifically, how three snippets of DBML JSON translate into the key elements of an ontology.

**Table 1.**
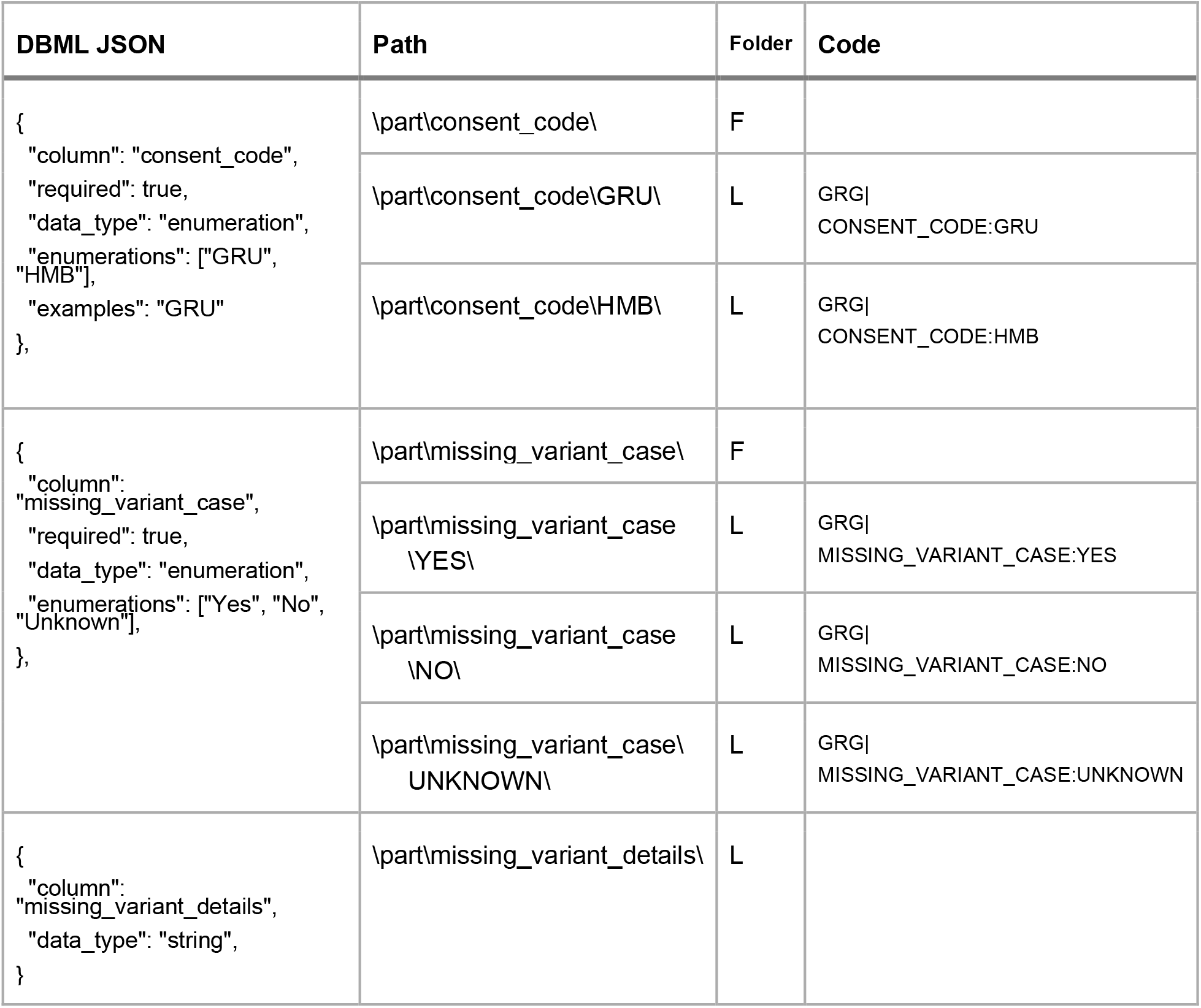
Portions of three DBML column definitions and their translation into i2b2 folders and leaves. The path defines a hierarchy, “folder” indicates whether the node is a (F)older or (L)eaf, and the code is a unique alphanumeric code identifying this path.

### Prompt: i2b2 facts

We next developed a prompt for conversion of a CSV data file conforming to the DBML dictionary into a pivoted one-row-per-fact i2b2 fact table. Our prompt first described the inputs: a data file to import, a mapping of patient identifiers from the data file into numeric pseudoidentifiers used by i2b2, and the ontology output by the previous prompt (for translating data values into i2b2 concept codes). Next, the prompt included technical documentation on i2b2 fact tables and ontology tables. Finally, a technical description of the approach in the Methods is presented.

Figure 4 shows an example of converting two rows of four fields into six i2b2 facts, using a patient_mapping table to translate from GREGoR identifiers to i2b2 pseudoidentifiers. For more information on the relationships between ontology and facts in data translation, see our earlier paper. [24]

**Figure 4.**
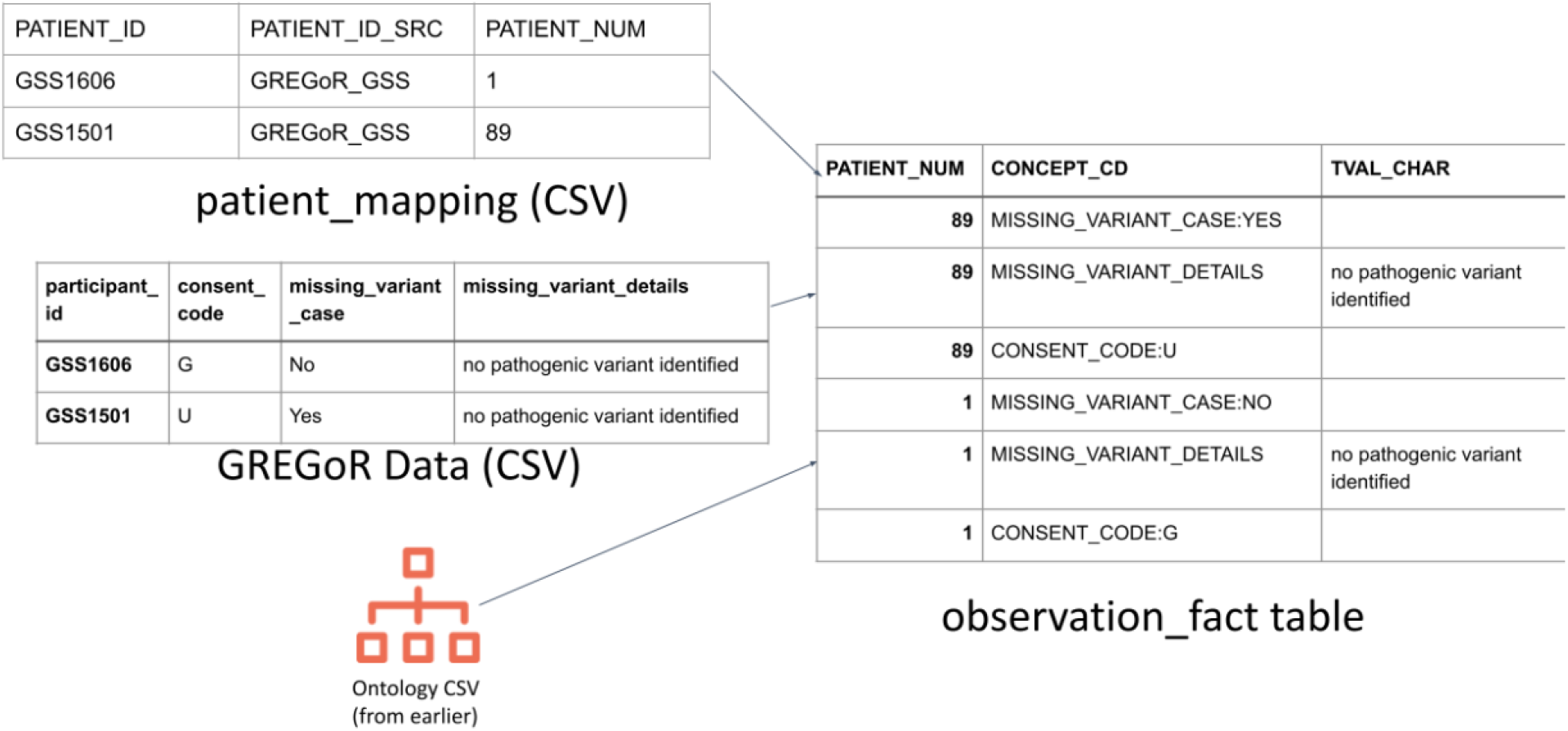
Example of running the generated Python script to convert GREGoR data rows into six i2b2 facts. GREGoR identifiers are converted to i2b2 pseudoidentifiers using an i2b2 patient_mapping table.

### Final data conversion

We executed our ChatGPT-4o-generated Python programs to convert the source files into i2b2 format, and we imported these to a SQL Server database connected to an i2b2 project and web interface. To support data encoded in the Human Phenotype (HP) and the Mondo Disease ontologies, terminologies not included in i2b2, we used the updated i2b2 NCBO Extractor. It accurately converted the BioPortal ontologies into i2b2 ontologies, building the entire hierarchy and placing unclassified concepts in a separate folder.

We used the standard i2b2 query tool to verify that the data loaded properly. The scripts correctly loaded data on 3458 participants’ data into ontologies for GREGoR Participant and Phenotype (94 participants), CMG Subject (1000 participants), eMerge 3 HRC Case/Control (2364 participants), HP (960 participants), Mondo (5 participants), and OMIM (139 participants). Figures 5 and 6 show a portion of these hierarchies.

**Figure 5.**
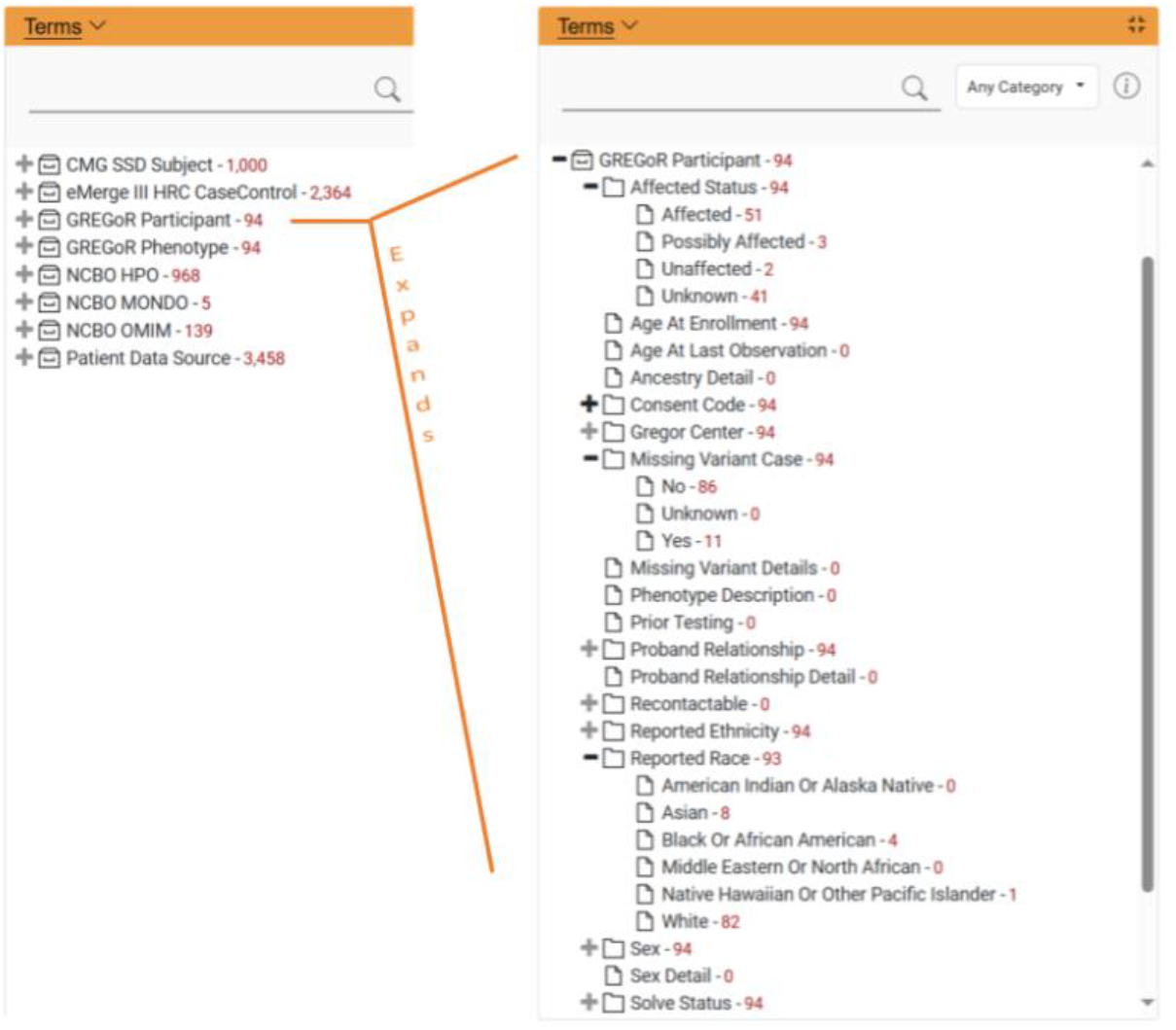
Two screenshots of loaded ontologies in our i2b2 webclient, showing ChatGPT-4o generated Python scripts successfully ETLed the GREGoR Participant ontology from its data dictionary and loaded its corresponding synthetic data. The left screen shows the top-level ontologies. Participant counts are in red. The numbers help verify the correctness of the Python scripts. On the right, the GREGoR Participant ontology is expanded to reveal the concepts and counts in multiple levels.

**Figure 6.**
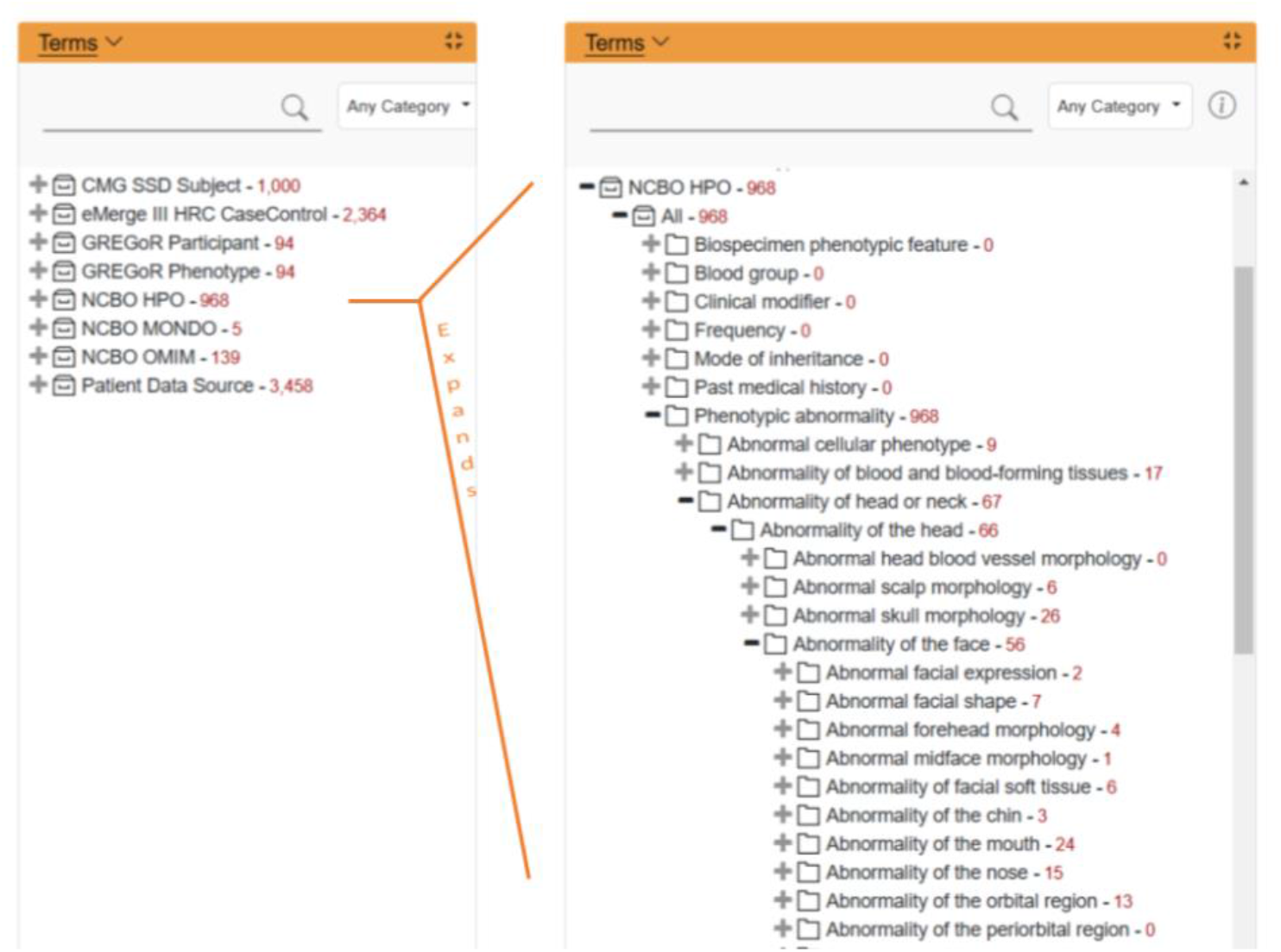
Two screenshots of loaded ontologies in our i2b2 webclient, showing the NCBO Human Phenotype Ontology (HPO) was successfully extracted from BioPortal and installed in i2b2. ChatGPT-4o generated Python scripts successfully ETLed corresponding synthetic participant data. The left screen shows the same top-level ontologies as Figure 5. On the right, the ontology tree is expanded to reveal the children of \NCBO HPO\Phenotypic abnormality\Abnormality of the head\Abnormality of the face\, showing the successful ETL of participant data to all levels of the ontology.

We validated the correctness of the data conversion in two ways. First, we examined the output of the Python program to verify that the program logic matched the functionality description in the prompt. Second, we computed patient counts for each ontology item using an automated script (note the numbers in Figure 5) and manually compared these to the input spreadsheets, using Excel functions to compute counts in the spreadsheets.

## Discussion

Developing new ETL processes for enabling spreadsheet data to be published into a multi-purpose database like i2b2 required many custom scripts for specific data models. This necessitated a skilled data scientist proficient in the source data model as well as i2b2 ontology design and data model. In contrast, we engineered a set of few-shot prompts and a pipeline utilizing GPT-4o to convert custom ontologies and their data into i2b2 via intermediary Python programs. All that is required for a new ETL process is to supply the technical descriptions of the data model and the steps for the conversion, in English text. This significantly reduces complexity and training requirements, enabling new types of data into i2b2.

Through this endeavor, we learned some important principles of prompt engineering:

- **Repeat important details several times throughout the prompt**. ChatGPT “forgets” about instructions given much earlier in a long prompt, because it is minimizing computational complexity by assuming that words near one another are more likely related than words that are distant. For example, the LLM frequently outputs programs allowing spaces in the C_FULLNAME, even when the prompt clearly indicates that spaces should be converted to underscores. The output program was correct after the prompt was modified to repeat this constraint close to a description of C_FULLNAME handling.
- **It is essential to have a programmer’s mindset when writing a prompt, as if writing high-level pseudocode**. Like a human, the LLM will make assumptions to fill in gaps if a detail isn’t specifically included. However, the LLM is more prone to making suboptimal (or even incorrect) decisions than an experienced programmer. Therefore prompt engineers must account for the consequences of ambiguity and try to cover the edge cases in the pseudocode.
- **A corollary to this: It is important to explicitly explain all the decision pathways in a conditional statement**. Explicit details are important. In a demographics merging program, the prompt explained that if a leaf node exists in two or more input ontologies, it should be replaced by a folder containing all of the leaves. However, the prompt did not explicitly explain what to do when a leaf existed in exactly one ontology. It did have a blanket statement that the input demographics should be unchanged if a case is unspecified, but the behavior of this was erratic. In one run, the output program removed the node entirely, while in another it created a folder with one leaf node. This was fixed when the prompt was modified to explicitly describe the missing case.
- **Let the LLM assist with debugging**. When the errors occurred in output program execution, it was frequently effective to paste the error into the ChatGPT prompt, which often was able to identify and correct the bug. A common issue involved missing type casting, where integers were being treated as integers in one context and character strings in another.
- Finally, **let the LLM inform decisions on structuring program requirements**, using iterative refinement and LLM-generated prompts. Iterative refinement was performed in response to unexpected output of the initial prompt. This was done by repeating the problematic portion of the prompt, explaining the problem, and requesting a corrected version of the code. Moreover, we could ask the LLM to update the prompt to clarify the issue. Starting with a slightly incomplete prompt was often advantageous, because it allowed the LLM to fill missing details into the prompt that could then be reviewed. For example, in one case we prompted the LLM to develop a Python program to persistently cache name lookups, and the LLM determined this was best done using the pickle.py object serialization module.

The potential time savings offered by this approach are substantial, but difficult to quantify precisely. The initial conversion of GREGoR into i2b2 took longer to do via LLM vs. manually, primarily because it required prompt-engineering methodology development and a library of reusable prompts and text components. However, converting the remaining data models took a matter of days instead of weeks. To the degree that the input structure resembled the output structure, new data conversions were largely trivial. The CMG data model aligned well with the logical structures of GREGoR’s DBML JSON, so only a data dictionary conversion prompt was needed. eMerge III required more effort. It specified a code for each value of an enumeration, which was not supported by GREGoR’s DBML. As a result, both the DBML specification and prompt text needed to be expanded.

A strong advantage of this approach over traditional ETL tools in i2b2 is its flexibility. It is frequently sufficient to modify only the prompt to support additional data models. i2b2-cdi, for example, supports creation of ontologies and facts in i2b2 by converting a particular spreadsheet format. [25] Our methodology enables conversion from arbitrary source formats without additional programming.

## Conclusion

Through carefully designed data mapping methodologies combined with biomedical ontologies and Large Language Models, we can provide easy-to-use tooling that will help bridge the gap between bespoke data models collected using spreadsheets and enterprise data warehouses like i2b2. Such tools will enable biomedical researchers using systems like AnVIL to utilize the power of i2b2 for cohort refinement and analytics.

## Data Availability

Prompts and Python code used in this study are in Github at https://github.com/jklann/LLMETL. The new NCBO Extraction Tool can be found at https://github.com/i2b2plugins/ncbo-extraction-tool. The synthetic data are available on request.

https://github.com/i2b2plugins/ncbo-extraction-tool

## Acknowledgements

We would like to acknowledge Robert Carroll and Anne Holmes at Vanderbilt. Robert recommended the three data models as good examples, and Anne generated the synthetic datasets. We also thank Jeremy Harper of Owl Health, whose insights into prompt engineering were invaluable in our methodology development.

## Author contributions

JGK conceived the study, designed the methodology, and led the development of the prompt**S**. SNM contributed to the study’s conceptual focus and refinement of its design. TDW developed the updated BioPortal–i2b2 ontology converter. VC assisted with prompt development and supported data conversion efforts. TDW and JGK drafted the manuscript, with all authors contributing to revisions and final edits.

